# Metabolomic/lipidomic profiling of COVID-19 and individual response to tocilizumab

**DOI:** 10.1101/2020.11.10.20228361

**Authors:** Gaia Meoni, Veronica Ghini, Laura Maggi, Alessia Vignoli, Alessio Mazzoni, Lorenzo Salvati, Manuela Capone, Anna Vanni, Leonardo Tenori, Paolo Fontanari, Federico Lavorini, Adriano Peris, Alessandro Bartoloni, Francesco Liotta, Lorenzo Cosmi, Claudio Luchinat, Francesco Annunziato, Paola Turano

## Abstract

The current pandemic emergence of novel coronavirus disease (COVID-19) poses a relevant threat to global health. SARS-CoV-2 infection is characterized by a wide range of clinical manifestations, ranging from absence of symptoms to severe forms that need intensive care treatment. Here, plasma-EDTA samples of 30 patients compared with age- and sex-matched controls were analyzed via untargeted nuclear magnetic resonance (NMR)-based metabolomics and lipidomics. With the same approach, the effect of tocilizumab administration was evaluated in a subset of patients. Despite the heterogeneity of the clinical symptoms, COVID-19 patients are characterized by common plasma metabolomic and lipidomic signatures (91.7% and 87.5% accuracy, respectively, when compared to controls). Tocilizumab treatment resulted in at least partial reversion of the metabolic alterations due to SARS-CoV-2 infection. In conclusion, NMR-based metabolomic and lipidomic profiling provides novel insights into the pathophysiological mechanism of human response to SARS-CoV-2 infection and to monitor treatment outcomes.

**Author summary:** The current COVID-19 pandemic caused by severe acute respiratory syndrome coronavirus-2 (SARS-CoV-2) is markedly affecting the world population. Here we report about the small-molecule profile of patients hospitalized during the first wave of the COVID-19 pandemic. Using magnetic resonance spectroscopy, we showed that the infection induces profound changes in the metabolome. The analysis of the specific metabolite changes and correlations with clinical data enabled the identification of potential biochemical determinants of the disease fingerprint. We also followed how metabolic alterations revert towards those of the control group upon treatment with tocilizumab, a recombinant humanised monoclonal antibody against the interleukin-6 receptor. These results open up possibilities for the monitoring of novel patients and their individual response to treatment.

## Introduction

The World Health Organization announced COVID-19 outbreak a pandemic in March 2020 [1,2]. At the beginning of October 2020 over thirty-four millions of patients have been diagnosed by SARS-CoV-2 infection and about 1 million deaths are reported all over the world [3]. The SARS-CoV-2 infection is characterized by a wide range of clinical manifestations, ranging from absence of symptoms to severe forms that need intensive care treatment. About 20% of patients, particularly the older ones and those affected by chronic comorbidities such as hypertension, diabetes mellitus, renal and heart diseases, may develop interstitial pneumonia and respiratory distress requiring oxygen therapy or mechanical ventilation [4]. In addition to interstitial pneumonia and acute respiratory distress syndrome (ARDS), COVID-19 is associated with other life-threatening complications such as sepsis, thromboembolism and multi-organ failure [5]. Patients with the highest rate of morbidity and mortality following SARS-CoV-2 infection develop a hyperinflammatory syndrome due to the overproduction of early response proinflammatory cytokines (such as IL-1β, IL-6, TNFα, MCP-1) – the so called “cytokine storm” – leading to an increased vascular permeability, activation of coagulation pathways, dysregulation of T cells with associated lymphopenia, multiorgan injury and rapid clinical deterioration [6–9].

Metabolomics and lipidomics can contribute a system level picture, thus expanding the options that chemists can explore to help fight the pandemic [10]. The human metabolome is composed by an ensemble of several thousands of small molecules (<1500–2000 Da) present on a very ample range of concentrations (from <1 nM to >1 μM) and produced by the genome of the host organism and by the genomes of its microflora, as well as deriving from exogenous factors like medical treatments [11]. Blood plasma is a primary carrier of small molecules in the body, the relative concentrations of which reflect the physio-pathological state of the organism and thus, possible tissue lesions and organ dysfunctions. The overall picture is complemented by alterations in the lipid components [12]. As a consequence, metabolomics and lipidomics of serum and plasma are increasingly used for successful patient stratification in various diseases [13–18]. Herein, a strong metabolomic and lipidomic signature of COVID-19 is revealed via untargeted nuclear magnetic resonance (NMR) spectroscopy of plasma-EDTA [19,20] from 30 patients compared with age- and sex-matched controls. Moreover, in a subset of patients, the metabolic effects due to tocilizumab administration were successfully investigated. This study had no sample-size calculation; the analysis included 30 patients who met the inclusion criteria and were admitted during the first wave of the pandemic at the local university hospital, before the rapid decline of hospitalizations for COVID-19.

## Results and Discussion

We analyzed via ^1^H NMR spectroscopy the metabolomic and lipidomic profiles of plasma-EDTA samples obtained from 30 patients affected by coronavirus disease 2019 (COVID-19). SARS-CoV-2 infection was confirmed by positive RT-PCR on nasopharyngeal swab specimens. The plasma-EDTA samples available for the metabolomic analysis were collected between 2-23 days from clinical onset (mean 9 days). Samples from 30 non COVID-19 subjects, one-to-one matched for age and sex, were used as control group (CTR). Tocilizumab, a humanized anti-IL-6 receptor monoclonal antibody, was administered to 8 of the 30 COVID-19 patients enrolled and another plasma-EDTA sample for each patient was collected after 2-18 days of treatment (mean 5 days). Demographic and clinical characteristics of enrolled patients are reported in S1 Table. Our analyses considered 21 quantified metabolites and 114 lipoprotein-related parameters [21]. Lipoprotein quantification of plasma samples of two COVID-19 patients (COVID-19-025 and COVID-19-027) was not possible for the presence of an interfering signal in the spectra, thus also their respective matched controls (CTR-4 and CTR-7) were removed from the lipoprotein analyses.

No outliers were identified using principal component analysis on the entire population, both for metabolite and lipoprotein profiles (S1 Fig).

Plasma metabolite and lipoprotein profiles of COVID-19 patients and CTRs were compared using the Random Forest (RF) algorithm. The eight samples collected post-tocilizumab treatment are not included in these analyses. The RF model built on metabolite concentrations shows a significant differential clustering with 91.7% accuracy, 93.3% sensitivity, and 90.0% specificity (Figs 1A and C, S2A Fig, Table S2). In particular, a panel of 11 metabolites (Fig 1D, S3 Table) displays significant alterations between COVID-19 patients and CTRs. One of them, giving rise to a detectable multiplet signal in the region between 7.21-7.30 ppm has not been assigned and is referred as “unknown”. However, even if this signal is removed from the statistical model, the discrimination accuracy between COVID-19 patients and CTRs does not change significantly.

**Fig 1.**
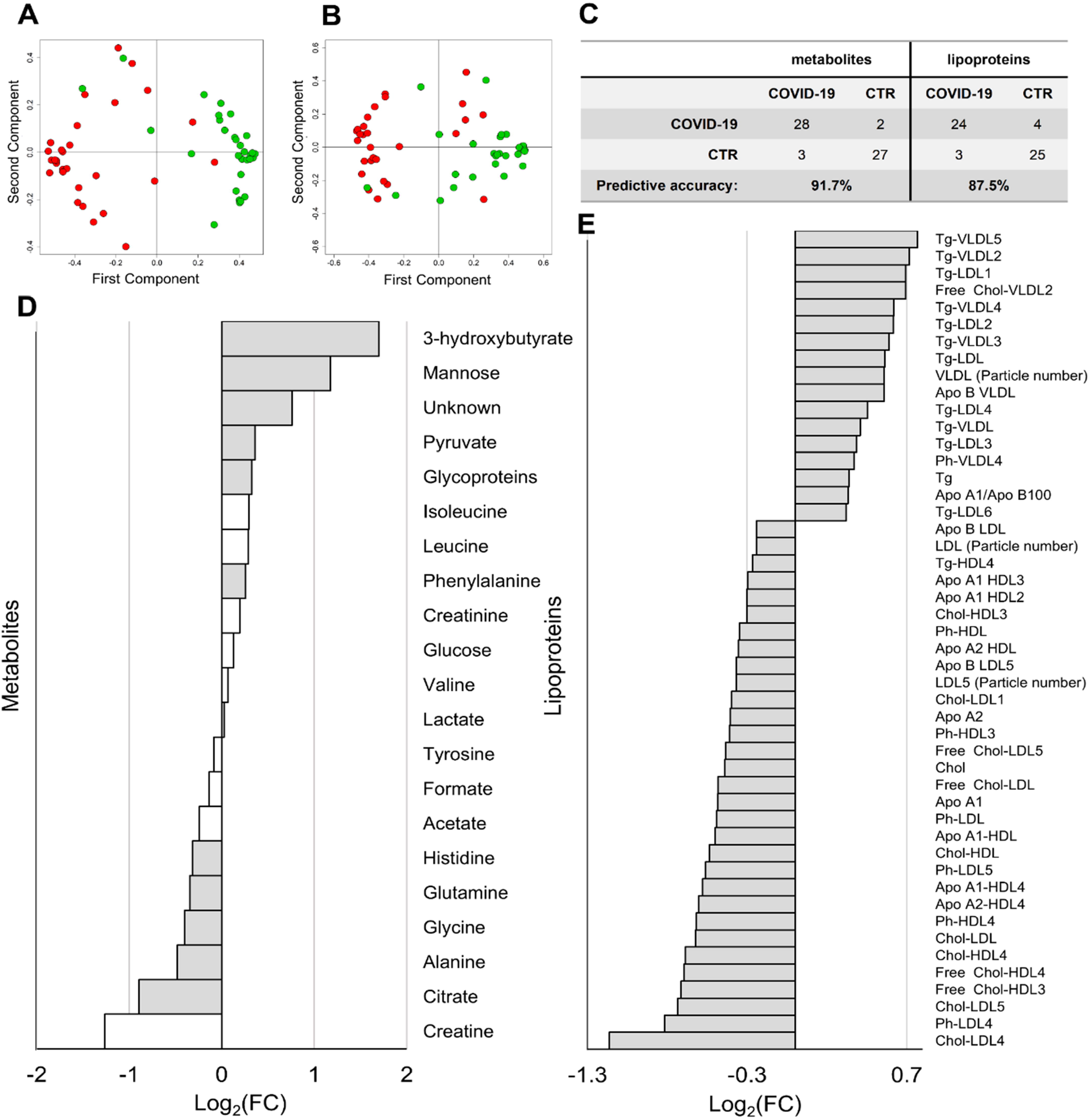
Metabolomic/lipidomic alterations in COVID-19 patients. (A-B) Proximity plots of the RF model discriminating COVID-19 patients (red dots), and CTR subjects (green dots) using A) the 21 quantified metabolites and B) the lipoprotein-related parameters. (C) Confusion matrices with predictive accuracy values of each model. (D) Values of Log_2_ (Fold Change, (FC) of quantified metabolites. Grey bars represent p-values < 0.05 after FDR correction. (E) Values of Log_2_(FC) of lipoprotein-related parameters significantly different (p-value < 0.05 after FDR correction) between COVID-19 patients and controls. Metabolites/lipoproteins with Log_2_(FC) positive/negative values have higher/lower concentration in plasma samples from COVID-19 patients with respect to controls.

The RF model built on lipoprotein-related parameters shows a significant differential clustering with 87.5% accuracy, 85.7% sensitivity, and 89.3% specificity (Fig 1B-C, S2B Fig, S2 Table). Fourty eight features (Fig 1E, S4 Table) display significant alterations between COVID-19 patients and CTRs. These results demonstrate that COVID-19 patients are characterized by higher levels of VLDL particles, and lower levels of Apo A1, Apo A2, cholesterol and free-cholesterol HDL and LDL subfractions. In particular, the subfractions HDL-3, HDL-4, LDL-4, LDL-5 of cholesterol are the most affected.

Correlations between clinical and metabolomic parameters were calculated and the results are reported in Fig 2.

**Fig 2.**
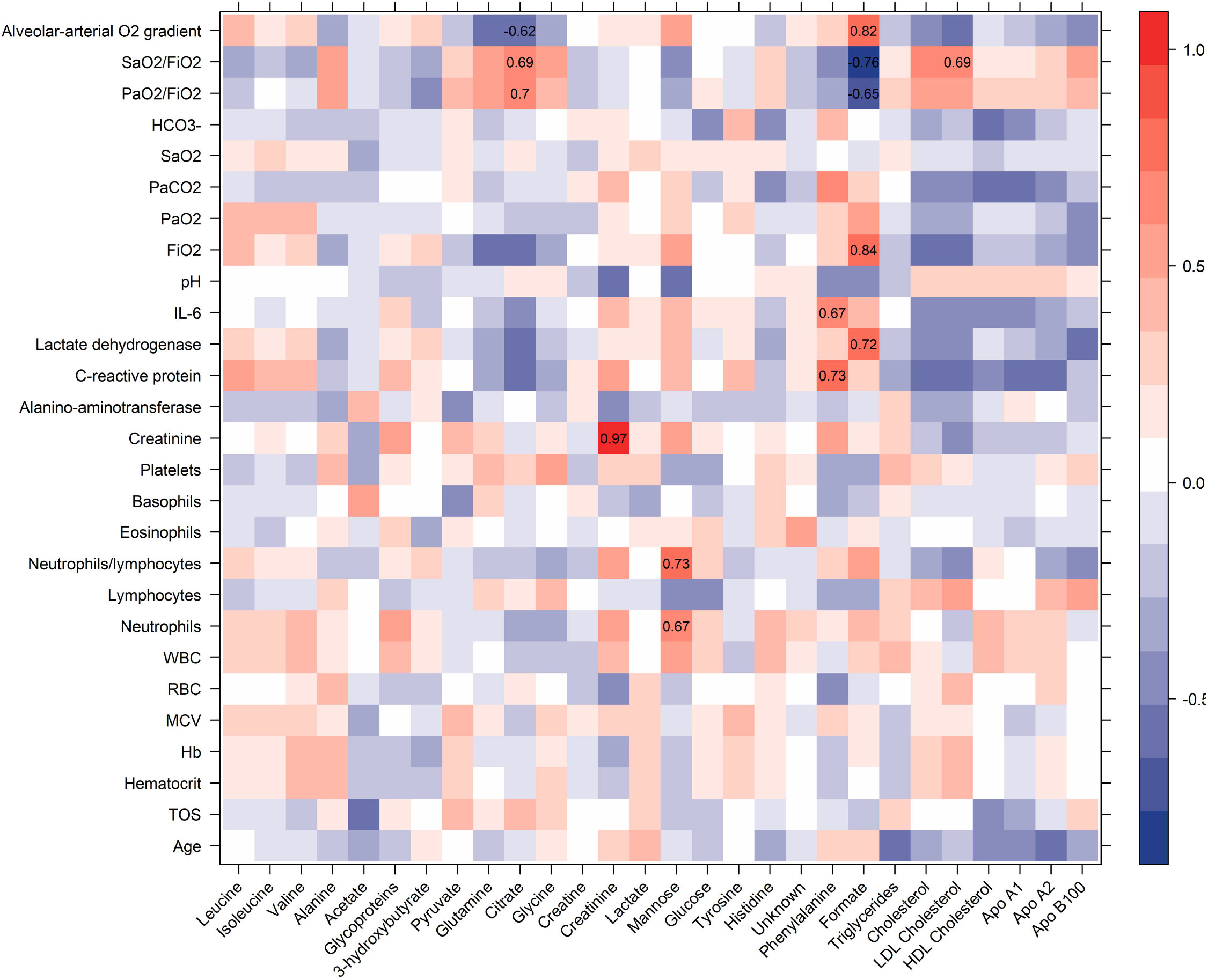
Heatmap correlations between clinical and metabolomic parameters. R values are shown as different degree of color intensity (red, positive correlations; blue, negative correlation). R values are reported in the plot only for statistically significant correlations (p-value < 0.05 after FDR correction).

Phenylalanine significantly correlates with C-reactive protein (CRP) and interleukin-6. Inflammation and immune activation impair the conversion of phenylalanine to tyrosine, as observed in patients suffering from sepsis, cancer, or HIV-1 infection [22–25]; accordingly, we found higher phenylalanine levels and a trend in lower tyrosine amounts in patients than in controls. Interestingly, a positive correlation between phenylalanine/tyrosine ratio and high CRP levels, has been already described by Murr and colleagues [26] in patients affected with coronary artery disease (CAD). These data are in accordance with ours, since SARS-CoV-2 infection is characterized not only by immune activation and systemic phlogosis, but also by microvascular endothelial damage and activation of coagulative cascade, as happens in CAD. Alveolar-arterial O_2_ gradient anticorrelates with citrate, accordingly the partial pressure of arterial oxygen and fraction of inspired oxygen (PaO_2_/FiO_2_) ratio (known as Horowitz index) and the ratio between oxygen saturation and fraction of inspired oxygen (SaO_2_/FiO_2_) positively correlates with citrate. This metabolite is known for its anti-oxidative, anti-coagulant and anti-inflammatory properties [27–29]. SARS-CoV-2 infection can cause lung damage, leading to ARDS, due not only to alveolar damage but also to diffuse microvascular endothelial damage and clot activation, mainly driven by pro-inflammatory cytokines, including IL-6 [30,31]. The protective role of citrate on endothelial integrity was recently reported by Dellepiane and colleagues [32]. Moreover, the consumption of citrate and other carboxylates is promoted by hypoxic conditions in red blood cells [33].

Formate shows the inverse pattern of correlation with respect to citrate, and it significantly correlates also with lactate dehydrogenase and FiO_2_. Regarding lipoprotein-related parameters only LDL cholesterol significantly correlates with the SaO_2_/FiO_2_ ratio.

In line with these observations, metabolites provide an optimal discrimination (accuracy 90.0%, 100.0% sensitivity, 83.3% specificity) between COVID-19 patients treated and non-treated with invasive ventilation (S3 A-B Fig), with formate and citrate as the most important features of the model. Instead, no significant clustering is present in the model calculated with lipoprotein-related parameters (S3 C-D Fig).

Despite plasma samples of COVID-19 patients are characterized by higher levels of VLDL and associated triglycerides, we observed a general reduction of HDL and LDL cholesterol-related parameters. Downregulation of lipids in COVID-19 blood sera has already been observed [34,35] and it has been hypothesized that lipids (in particular cholesterol and fatty acids) could play a pivotal role in virus replication and assembly [36,37]. Our data suggest that only LDL and HDL could be implied in this mechanism.

Accordingly, in a recent study, LDL and HDL levels were inversely correlated to disease severity and poor prognosis [38]. Furthermore, overproduction of VLDL has been linked with the processes inducing insulin resistance in COVID-19 patients [35].

We also detected an accumulation of mannose in the plasma of COVID-19 patients and a significant positive correlation between plasma mannose levels and neutrophils and between mannose and the neutrophils to lymphocytes ratio. An increment of mannose could be related to different reasons: it could be associated to its binding to lectin in order to promote complement activation [34], or it could be linked to insulin resistance. Indeed, plasma mannose levels are elevated in subjects with insulin resistance independently of obesity [39] and there are increasing evidences that a bidirectional relationship between COVID-19 and diabetes exists [40].

The increment of pyruvate and 3-hydroxybutyrate, along with the strong decrement of citrate and free amino acids (alanine, glycine, glutamine, histidine) in plasma of COVID-19 patients can be ascribed to an impairment of the energetic metabolism. Indeed, during inflammatory states amino acids can be used to provide energy and materials for the proliferation and phagocytosis of immune cells. It is important to underline that pyruvate is a metabolite particularly sensitive to pre-analytical procedures, thus further investigations to confirm its alteration are needed [41,42].

Among the 30 COVID-19 patients enrolled, 18 patients presented ARDS. Thus, the possibility that ARDS could significantly alter the profile of COVID-19 was examined. The RF models calculated both on metabolites and lipoprotein-related parameters can only slightly cluster ARDS patients with respect to the other COVID-19 patients (metabolite model: accuracy 76.7%, 88.9% sensitivity, 58.3% specificity; lipoprotein model: accuracy 75.0%, 81.2% sensitivity, 66.7% specificity). These results demonstrate that the metabolomic profile of COVID-19 patients is mainly dictated by the pathology or by the host response to the virus infection, rather than by the concomitant presence of ARDS.

Multilevel partial least square discriminant analysis (mPLS-DA) was used to analyze NMR data of pre- and post-tocilizumab samples in a pairwise multivariate fashion. The mPLS-DA model built on metabolites shows significant differential clustering, yielding a discrimination accuracy of 80.3% (Fig 3A). The two pairs of samples collected from patients who died (COVID-19-020 and COVID-19-021) present the smallest shift within the metabolomic subspace. Unfortunately, COVD-19-018 patient was transferred to another hospital and no follow-up and outcome information was available. Univariate analysis enables the identification of a panel of eight metabolites (Fig 3B-I, S5 Table) significantly different (before FDR correction) between pre- and post-tocilizumab patients. The post-treatment levels of these metabolites partially or completely revert towards the levels of CTR subjects.

**Fig 3.**
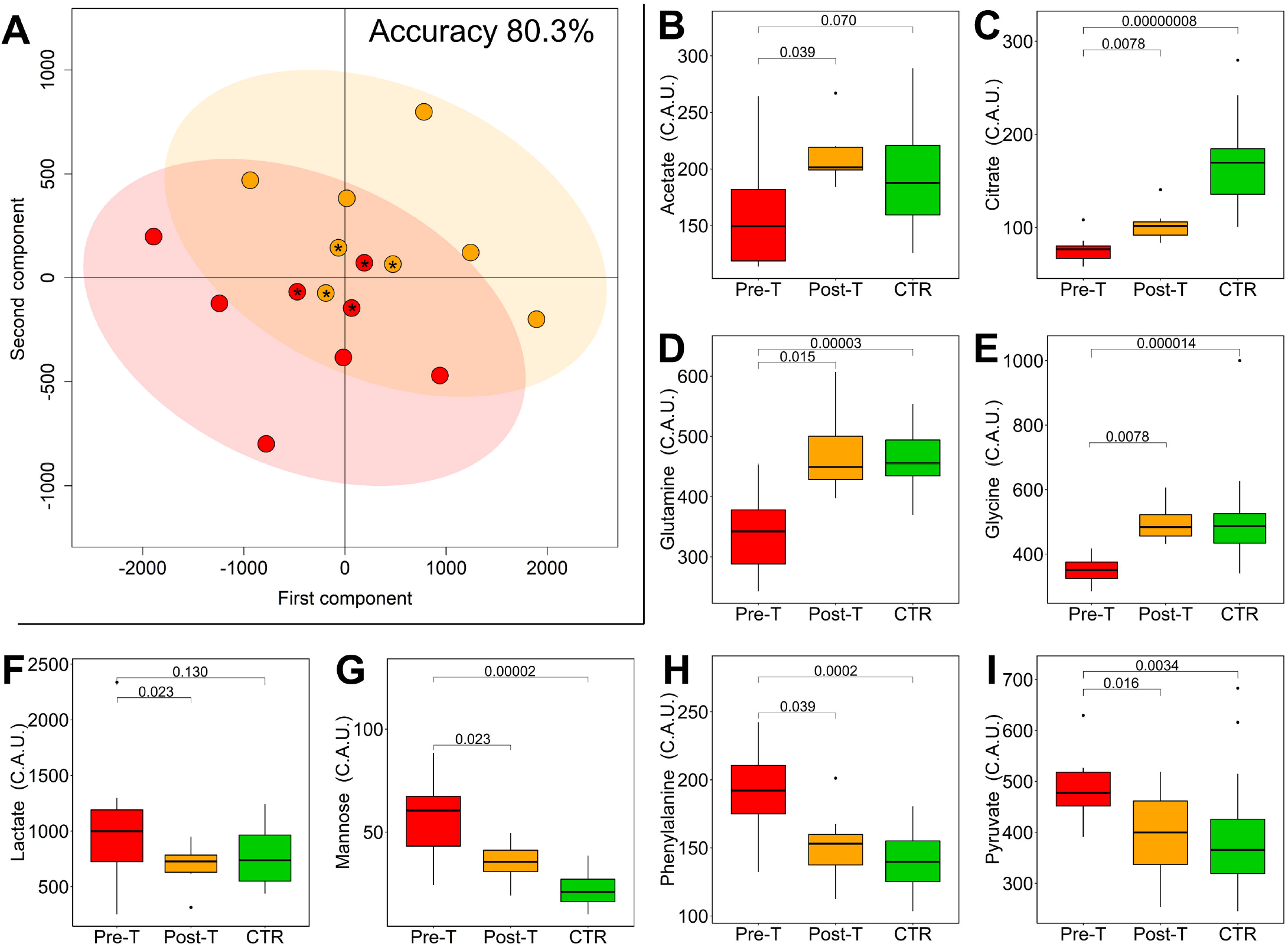
Tocilizumab treatment reverts metabolomic/lipidomic alterations in COVID-19 patients. (A) Score plot (of the first two principal components) and accuracy of the mPLS-DA model discriminating COVID-19 patients at pre- (red dots) and post- (orange dots) tocilizumab treatment using the 21 quantified metabolites. The code of each patient is reported inside each dot. Patients 20 and 21 died. (B-I) Boxplots of the statistically significant metabolites discriminating of pre- (red) and post- (orange) tocilizumab samples, p-values obtained using Wilcoxon signed-rank test are also reported. Boxplots of controls (green) and the p-values (Wilcoxon-Mann-Whitney test) for the comparison between pre-treatment and CTR are reported. P-values adjusted for FDR are reported in Table S5.

The mPLS-DA model built using lipoprotein-related parameters shows a significant discrimination between samples collected pre- and post-tocilizumab treatment (accuracy of 82.8%) (S4 Fig). Univariate analysis identifies 19 lipoprotein parameters (Table S6) significantly different between pre- and post-tocilizumab treatment. In particular, HDL-1 subfractions of cholesterol, phospholipids, and Apo A2 showed lower levels at post-treatment, whereas LDL-5, HDL-4, IDL, VLDL-1, and VLDL-2 of many subfractions are higher at post-treatment. The general increment of lipoprotein subfractions after treatment confirms the metabolic reversion and supports the key role of lipids in the metabolism of COVID-19 patients.

In summary, in this study a strong plasma metabolomic and lipidomic signature of Sars-CoV-2 infection is identified, and it is only marginally affected by the different disease severities. Additionally, some metabolites correlate with alveolar-capillary membrane injury and their levels are affected by mechanical ventilation. Of note, in the case of patients who underwent tocilizumab treatment, metabolic alterations revert towards those of the control group.

## Materials and Methods

### Study approval

The study was approved by the Comitato Etico Regionale-sezione area vasta centro (protocol 16859) and it complies with the 1964 Declaration of Helsinki and its later amendments. Written informed consent was obtained from recruited patients.

### Patients characteristics

During the first wave of SARS-CoV-2 infection, 30 patients, hospitalized at the Careggi University Hospital, Florence, Italy, were enrolled in the study. All patients were of Caucasian ethnicity. Demographic and clinical features of the enrolled patients are summarized in Table S1.

### Plasma sample preparation

Plasma samples were collected from all the subjects enrolled in the study, according to standard procedures [41,42]. Blood samples were collected in ethylene diamine tetra-acetate (EDTA)-coated blood collection tubes and stored at room temperature. Ficoll®, a neutral highly branched polymer formed by the co-polymerization of sucrose and epichlorohydrin, was used for blood separation. 14 mL of blood were gently placed in 25 mL tubes containing 9 mL of Ficoll®. Tubes were centrifugated 1500 g for 20 minutes. Plasma was collected and rapidly stored in a −20°C freezer pending NMR analysis.

### NMR sample preparation, spectra processing and spectral analysis

NMR samples were prepared according to standard procedures [19,20,43]. NMR spectra for all samples were acquired using a Bruker 600 MHz spectrometer (Bruker BioSpin) operating at 600.13 MHz proton Larmor frequency and equipped with a 5 mm PATXI ^1^H-^13^C-^15^N and ^2^H-decoupling probe including a z-axis gradient coil, an automatic tuning-matching (ATM) and an automatic refrigerated sample changer (SampleJet, Bruker BioSpin). A BTO 2000 thermocouple served for temperature stabilization at the level of approximately 0.1 K of the sample. Before measurement, to equilibrate temperature at 310 K, samples were kept for at least 5 minutes inside the NMR probe head.

For each plasma sample, two one-dimensional ^1^H NMR spectra were acquired with water peak suppression and different pulse sequences that allowed the selective detection of different molecular components. The spectra were: 1) a standard NOESY using 32 scans, 98,304 data points, a spectral width of 18,028 Hz, an acquisition time of 2.7 s, a relaxation delay of 4 s and a mixing time of 0.01 s. This kind of spectrum is made up of signals arising from low molecular weight molecules (metabolites) and signals arising from macromolecules such as lipoproteins and lipids; 2) a standard spin echo Carr-Purcell-Meiboom-Gill (CPMG) using 32 scans, 73,728 data points, a spectral width of 12,019 Hz and a relaxation delay of 4 s. This NMR sequence allows the selective detection of signals arising only from low molecular weight metabolites.

Before applying Fourier transform, free induction decays were multiplied by an exponential function equivalent to a 0.3 Hz line-broadening factor. Transformed spectra were automatically corrected for phase and baseline distortions and calibrated to the anomeric glucose doubled at δ 5.24 ppm, using TopSpin 3.6.2 (Bruker BioSpin) [19,20].

### Statistical analysis

All data analyses were performed using the “R” statistical environment. Metabolites, whose peaks in the CPMG spectra were well defined and resolved, were assigned and their concentrations analyzed. The assignment procedure was performed using an ^1^H NMR spectra library of pure organic compounds (BBIOREFCODE, Bruker BioSpin), public databases, *e.g*. Human Metabolome Database [11], storing reference ^1^H NMR spectra of metabolites, and using literature data when available. The spectral regions associated to the 21 assigned metabolites were integrated using a R script in-house developed. Quantification of 114 lipid main fractions and subfractions was performed using the Bruker IVDr platform [21].

Both metabolites and lipoprotein-related parameters were analyzed via multivariate analysis. Unsupervised Principal Component Analysis (PCA) was used as first exploratory analysis to obtain a preliminary data outlook (*i.e*. cluster detection and screening for outliers). The Random Forest (RF) algorithm [44] was used for classification in the comparison between COVID-19 patients and CTR. RF is a classification algorithm that uses an ensemble of unpruned decision trees (forest), each of which is built on a bootstrap sample of the training data using a randomly selected subset of variables (metabolites or lipoprotein-related parameters)[45,46]. The percentage of trees in the forest that assign one sample to a specific class can be inferred as a probability of belonging to a given class [13]. In our case, each tree was used to predict whether a sample represents a COVID-19 patient or a CTR subject. Because the out-of-bag (OOB) observations were not used in the fitting of the trees, the OOB estimates are cross-validated, accuracy estimates, and represent an unbiased estimation of the generalization error [44]. Accuracy, sensitivity, and specificity of all calculated models were assessed according to the standard definitions. For all calculations, the R package ‘Random Forest’ [44] was used to grow a forest of 1000 trees, using the default settings.

Pairwise comparisons between pre- and post-treatment samples were performed using multilevel Partial Least Square Discriminant Analysis (mPLS-DA) and results validated using a Monte Carlo Cross-Validation scheme (MCCV, script in house developed): each dataset was randomly divided by 1000 times into a training set (80% of the data) which was used to build the model and a test set (20% of the data) which was used to test the integrity of the model. Accuracy, sensitivity, and specificity of all calculated models were assessed according to the standard definitions.

On the biological assumption that metabolite and lipoprotein concentrations are not normally distributed, non-parametric tests were used for the univariate analysis. Wilcoxon-Mann-Whitney test was used to infer differences between the metabolite/lipid levels in the comparison between COVID-19 patients and CTR. Instead for pairwise comparison between pre- and post-treatment samples the Wilcoxon signed-rank test was utilized [47]. P-values were adjusted for multiple testing using the false discovery rate (FDR) procedure with Benjamini-Hochberg [48] correction at α = 0.05. The effect size (Ef) was also calculated [49] to aid in the identification of the meaningful signals giving an estimation of the magnitude of the separation between the different groups. The magnitude is assessed using the thresholds provided in Romano et al. [50], *i.e*. |Ef|□<□0.147 “negligible”, |Ef|□<□0.33 “small”, |Ef|□<□0.474 “medium”, otherwise “large”. Pearson correlations, adjusted for FDR using BH methods, were also calculated.

## Data Availability

All the NMR files will be available from the MetaboLights database after publication.

## Acknowledgments

P.T., C.L., L.T., V.G., A.V. and G.M. acknowledge the support and the use of resources of Instruct-ERIC, a Landmark ESFRI project, and specifically the CERM/CIRMMP Italy Centre. AV and VG are supported by an AIRC fellowship for Italy. This study was supported by funds from the Department of Experimental and Clinical Medicine of University of Florence, “Excellence Departments 2018–2022 Project”.

## Supporting information

**S1 Fig. PCA analysis**. Score plots (PC1 vs. PC2) of the unsupervised PCA model of A) 21 quantified metabolites, B) lipoprotein-related parameters; COVID-19 patients (red dots); CTR subjects (green dots).

**S2 Fig. Metabolomic/lipidomic alterations in COVID-19 patients**. Variable importance plots of the Random Forest models discriminating COVID-19 patients and control subjects. A) 21 quantified metabolites, B) lipoprotein-related parameters.

**S3 Fig. Alterations induced by invasive ventilation in COVID-19 patients**. Proximity plot (of the first two principal components) and accuracy of the Random Forest model discriminating COVID-19 patients treated (blue dots) and non-treated (sea green dots) with invasive ventilation using metabolites (A) and lipoprotein-related parameters (C). Variable importance plots of the two Random Forest models: B) 21 quantified metabolites, D) lipoprotein-related parameters.

**S4 Fig. Alterations in lipoprotein profile induced by Tocilizumab treatment**. A) Score plot (of the first two principal components) and accuracy of the mPLS-DA model discriminating COVID-19 patients pre- (red dots) and post- (orange dots) tocilizumab treatment using the lipoprotein-related parameters.

**S1 Table. Demographic and clinical characteristics of COVID-19 patients**. ACF denotes acute cardiac failure, AI autoimmune disease, AKI acute kidney injury, ARDS acute respiratory distress syndrome, CKD chronic kidney disease, CVD cardiovascular disease, D deceased, DBT type 2 diabetes, DH discharged home, DYS dyslipidemia, H hypertension, K cancer, LTFU lost to follow-up.

**S2 Table. Metabolomic/lipidomic alterations in COVID-19 patients: multivariate analysis**. Random Forest scores of the model discriminating COVID-19 patients and controls using the 21 quantified metabolites and lipoprotein-related parameters. P: predicted class; S. numeric score (controls: 0<S< 0.5; COVID-19 patients: 0.5 < S< 1).

**S3 Table. Metabolomic alterations in COVID-19 patients: univariate analysis**. Univariate analysis of the 21 quantified metabolites for the comparison between COVID-19 patients and control subjects. The median and MAD of each metabolite in the two groups are reported. The p-value of the univariate Wilcoxon-Mann-Whitney test together with the p-value calculated after false discovery rate correction and the effect size, using the Cliff’s delta formulation, were also reported for each metabolite.

**S4 Table. Lipidomic alterations in COVID-19 patients: univariate analysis**. Univariate analysis of the lipoprotein-related parameters for the comparison between COVID-19 patients and control subjects. The median and MAD of each parameter in the two groups are reported. The p-value of the univariate Wilcoxon-Mann-Whitney test together with the p-value calculated after false discovery rate correction and the effect size, using the Cliff’s delta formulation, were also reported for each parameter.

**S5 Table. Metabolomic alterations induced by Tocilizumab treatment: univariate analysis**. Univariate analysis of the 21 quantified metabolites for the comparison between COVID-19 patients before and after tocilizumab treatment. The p-value of the univariate Wilcoxon-Signed-Rack test together with the p-value calculated after false discovery rate correction and the effect size were reported for each metabolite.

**S6 Table. Lipidomic alterations induced by Tocilizumab treatment: univariate analysis**. Univariate analysis of the lipoprotein-related parameters for the comparison between COVID-19 patients before and after tocilizumab treatment. The p-value of the univariate Wilcoxon-Signed-Rack test together with the p-value calculated after false discovery rate correction and the effect size were reported for each parameter.

## Notes

### Competing Interest Statement

The authors have declared no competing interest.

### Funding Statement

Unfunded study

## References

1. WHO Director-General’s opening remarks at the media briefing on COVID-19 - 2 October 2020. [cited 9 Oct 2020]. Available: <<https://www.who.int/dg/speeches/detail/who-director-general-s-opening-remarks-at-the-media-briefing-on-covid-192-october-2020>>

2. Zhou P, Yang X-L, Wang X-G, Hu B, Zhang L, Zhang W, et al. A pneumonia outbreak associated with a new coronavirus of probable bat origin. Nature. 2020;579: 270–273. doi:10.1038/s41586-020-2012-7

3. Coronavirus Disease (COVID-19) Situation Reports. [cited 9 Oct 2020]. Available: <<https://www.who.int/emergencies/diseases/novel-coronavirus-2019/situation-reports>>

4. Wu Z, McGoogan JM. Characteristics of and Important Lessons From the Coronavirus Disease 2019 (COVID-19) Outbreak in China: Summary of a Report of 72D314 Cases From the Chinese Center for Disease Control and Prevention. JAMA. 2020;323: 1239–1242. doi:10.1001/jama.2020.2648

5. Zhou F, Yu T, Du R, Fan G, Liu Y, Liu Z, et al. Clinical course and risk factors for mortality of adult inpatients with COVID-19 in Wuhan, China: a retrospective cohort study. Lancet (London, England). 2020;395: 1054–1062. doi:10.1016/S0140-6736(20)30566-3

6. Costela-Ruiz VJ, Illescas-Montes R, Puerta-Puerta JM, Ruiz C, Melguizo-Rodríguez L. SARS-CoV-2 infection: The role of cytokines in COVID-19 disease. Cytokine & Growth Factor Reviews. 2020;54: 62–75. doi:10.1016/j.cytogfr.2020.06.001

7. Mazzoni A, Salvati L, Maggi L, Capone M, Vanni A, Spinicci M, et al. Impaired immune cell cytotoxicity in severe COVID-19 is IL-6 dependent. The Journal of Clinical Investigation. 2020;130: 4694–4703. doi:10.1172/JCI138554

8. Vultaggio A, Vivarelli E, Virgili G, Lucenteforte E, Bartoloni A, Nozzoli C, et al. Prompt Predicting of Early Clinical Deterioration of Moderate-to-Severe COVID-19 Patients: Usefulness of a Combined Score Using IL-6 in a Preliminary Study. J Allergy Clin Immunol Pract. 2020;8: 2575-2581.e2. doi:10.1016/j.jaip.2020.06.013

9. Huang C, Wang Y, Li X, Ren L, Zhao J, Hu Y, et al. Clinical features of patients infected with 2019 novel coronavirus in Wuhan, China. The Lancet. 2020;395: 497–506. doi:10.1016/S0140-6736(20)30183-5

10. Opatz T, Senn-Bilfinger J, Richert C. Thoughts on What Chemists Can Contribute to Fighting SARS-CoV-2 – A Short Note on Hand Sanitizers, Drug Candidates and Outreach. Angew Chem Int Ed Engl. 2020;59: 9236–9240. doi:10.1002/anie.202004721

11. Wishart DS, Feunang YD, Marcu A, Guo AC, Liang K, Vázquez-Fresno R, et al. HMDB 4.0: the human metabolome database for 2018. Nucleic Acids Research. 2018;46: D608–D617. doi:10.1093/nar/gkx1089

12. Yang K, Han X. Lipidomics: Techniques, Applications, and Outcomes Related to Biomedical Sciences. Trends in Biochemical Sciences. 2016;41: 954–969. doi:10.1016/j.tibs.2016.08.010

13. Vignoli A, Tenori L, Giusti B, Takis PG, Valente S, Carrabba N, et al. NMR-based metabolomics identifies patients at high risk of death within two years after acute myocardial infarction in the AMI-Florence II cohort. BMC medicine. 2019;17: 3. doi:10.1186/s12916-018-1240-2

14. Meoni G, Lorini S, Monti M, Madia F, Corti G, Luchinat C, et al. The metabolic fingerprints of HCV and HBV infections studied by Nuclear Magnetic Resonance Spectroscopy. Scientific Reports. 2019;9: 4128. doi:10.1038/s41598-019-40028-4

15. McCartney A, Vignoli A, Biganzoli L, Love R, Tenori L, Luchinat C, et al. Metabolomics in breast cancer: A decade in review. Cancer Treatment Reviews. 2018;67: 88–96. doi:10.1016/j.ctrv.2018.04.012

16. Monnerie S, Comte B, Ziegler D, Morais JA, Pujos-Guillot E, Gaudreau P. Metabolomic and Lipidomic Signatures of Metabolic Syndrome and its Physiological Components in Adults: A Systematic Review. Scientific Reports. 2020;10: 669. doi:10.1038/s41598-019-56909-7

17. Bertini I, Cacciatore S, Jensen BV, Schou JV, Johansen JS, Kruhøffer M, et al. Metabolomic NMR Fingerprinting to Identify and Predict Survival of Patients with Metastatic Colorectal Cancer. Cancer Res. 2012;72: 356–364. doi:10.1158/0008-5472.CAN-11-1543

18. Zhang L, Zhu B, Zeng Y, Shen H, Zhang J, Wang X. Clinical lipidomics in understanding of lung cancer: Opportunity and challenge. Cancer Letters. 2020;470: 75–83. doi:10.1016/j.canlet.2019.08.014

19. Vignoli A, Ghini V, Meoni G, Licari C, Takis PG, Tenori L, et al. High-Throughput Metabolomics by 1D NMR. Angew Chem Int Ed Engl. 2019;58: 968–994. doi:10.1002/anie.201804736

20. Takis PG, Ghini V, Tenori L, Turano P, Luchinat C. Uniqueness of the NMR approach to metabolomics. TrAC Trends in Analytical Chemistry. 2019;120: 115300. doi:10.1016/j.trac.2018.10.036

21. Jiménez B, Holmes E, Heude C, Tolson RF, Harvey N, Lodge SL, et al. Quantitative Lipoprotein Subclass and Low Molecular Weight Metabolite Analysis in Human Serum and Plasma by 1H NMR Spectroscopy in a Multilaboratory Trial. Anal Chem. 2018;90: 11962–11971. doi:10.1021/acs.analchem.8b02412

22. Neurauter G, Schröcksnadel K, Scholl-Bürgi S, Sperner-Unterweger B, Schubert C, Ledochowski M, et al. Chronic immune stimulation correlates with reduced phenylalanine turnover. Curr Drug Metab. 2008;9: 622–627. doi:10.2174/138920008785821738

23. Ploder M, Neurauter G, Spittler A, Schroecksnadel K, Roth E, Fuchs D. Serum phenylalanine in patients post trauma and with sepsis correlate to neopterin concentrations. Amino Acids. 2008;35: 303–307. doi:10.1007/s00726-007-0625-x

24. Zangerle R, Kurz K, Neurauter G, Kitchen M, Sarcletti M, Fuchs D. Increased blood phenylalanine to tyrosine ratio in HIV-1 infection and correction following effective antiretroviral therapy. Brain Behav Immun. 2010;24: 403–408. doi:10.1016/j.bbi.2009.11.004

25. Capuron L, Schroecksnadel S, Féart C, Aubert A, Higueret D, Barberger-Gateau P, et al. Chronic low-grade inflammation in elderly persons is associated with altered tryptophan and tyrosine metabolism: role in neuropsychiatric symptoms. Biol Psychiatry. 2011;70: 175–182. doi:10.1016/j.biopsych.2010.12.006

26. Murr C, Grammer TB, Meinitzer A, Kleber ME, März W, Fuchs D. Immune activation and inflammation in patients with cardiovascular disease are associated with higher phenylalanine to tyrosine ratios: the ludwigshafen risk and cardiovascular health study. J Amino Acids. 2014;2014: 783730. doi:10.1155/2014/783730

27. Grundström G, Christensson A, Alquist M, Nilsson L-G, Segelmark M. Replacement of acetate with citrate in dialysis fluid: a randomized clinical trial of short term safety and fluid biocompatibility. BMC Nephrol. 2013;14: 216. doi:10.1186/1471-2369-14-216

28. Gabutti L, Lucchini B, Marone C, Alberio L, Burnier M. Citrate-vs. acetate-based dialysate in bicarbonate haemodialysis: consequences on haemodynamics, coagulation, acid-base status, and electrolytes. BMC Nephrol. 2009;10: 7. doi:10.1186/1471-2369-10-7

29. Marengo M, Dellepiane S, Cantaluppi V. Extracorporeal Treatments in Patients with Acute Kidney Injury and Sepsis. Contrib Nephrol. 2017;190: 1–18. doi:10.1159/000468912

30. Cipolloni L, Sessa F, Bertozzi G, Baldari B, Cantatore S, Testi R, et al. Preliminary Post-Mortem COVID-19 Evidence of Endothelial Injury and Factor VIII Hyperexpression. Diagnostics (Basel). 2020;10. doi:10.3390/diagnostics10080575

31. Lai C-C, Shih T-P, Ko W-C, Tang H-J, Hsueh P-R. Severe acute respiratory syndrome coronavirus 2 (SARS-CoV-2) and coronavirus disease-2019 (COVID-19): The epidemic and the challenges. Int J Antimicrob Agents. 2020;55: 105924. doi:10.1016/j.ijantimicag.2020.105924

32. Dellepiane S, Medica D, Guarena C, Musso T, Quercia AD, Leonardi G, et al. Citrate anion improves chronic dialysis efficacy, reduces systemic inflammation and prevents Chemerin-mediated microvascular injury. Scientific Reports. 2019;9: 10622. doi:10.1038/s41598-019-47040-8

33. Nemkov T, Sun K, Reisz JA, Yoshida T, Dunham A, Wen EY, et al. Metabolism of Citrate and Other Carboxylic Acids in Erythrocytes As a Function of Oxygen Saturation and Refrigerated Storage. Front Med. 2017;4. doi:10.3389/fmed.2017.00175

34. Shen B, Yi X, Sun Y, Bi X, Du J, Zhang C, et al. Proteomic and Metabolomic Characterization of COVID-19 Patient Sera. Cell. 2020;182: 59-72.e15. doi:10.1016/j.cell.2020.05.032

35. Kimhofer T, Lodge S, Whiley L, Gray N, Loo RL, Lawler NG, et al. Integrative Modeling of Quantitative Plasma Lipoprotein, Metabolic, and Amino Acid Data Reveals a Multiorgan Pathological Signature of SARS-CoV-2 Infection. J Proteome Res. 2020 [cited 12 Oct 2020]. doi:10.1021/acs.jproteome.0c00519

36. Schoggins JW, Randall G. Lipids in Innate Antiviral Defense. Cell Host Microbe. 2013;14: 379–385. doi:10.1016/j.chom.2013.09.010

37. Abu-Farha M, Thanaraj TA, Qaddoumi MG, Hashem A, Abubaker J, Al-Mulla F. The Role of Lipid Metabolism in COVID-19 Virus Infection and as a Drug Target. Int J Mol Sci. 2020;21. doi:10.3390/ijms21103544

38. Fan J, Wang H, Ye G, Cao X, Xu X, Tan W, et al. Letter to the Editor: Low-density lipoprotein is a potential predictor of poor prognosis in patients with coronavirus disease 2019. Metabolism. 2020;107: 154243. doi:10.1016/j.metabol.2020.154243

39. Mardinoglu A, Stančáková A, Lotta LA, Kuusisto J, Boren J, Blüher M, et al. Plasma Mannose Levels Are Associated with Incident Type 2 Diabetes and Cardiovascular Disease. Cell Metabolism. 2017;26: 281–283. doi:10.1016/j.cmet.2017.07.006

40. Rubino F, Amiel SA, Zimmet P, Alberti G, Bornstein S, Eckel RH, et al. New-Onset Diabetes in Covid-19. New England Journal of Medicine. 2020;383: 789–790. doi:10.1056/NEJMc2018688

41. Bernini P, Bertini I, Luchinat C, Nincheri P, Staderini S, Turano P. Standard operating procedures for pre-analytical handling of blood and urine for metabolomic studies and biobanks. J Biomol NMR. 2011;49: 231–243. doi:10.1007/s10858-011-9489-1

42. Ghini V, Quaglio D, Luchinat C, Turano P. NMR for sample quality assessment in metabolomics. N Biotechnol. 2019;52: 25–34. doi:10.1016/j.nbt.2019.04.004

43. PD CEN/TS 16945:2016 - Molecular in vitro diagnostic examinations. Specifications for pre-examination processes for metabolomics in urine, venous blood serum and plasma. [cited 12 Oct 2020]. Available: https://shop.bsigroup.com/ProductDetail?pid=000000000030339067

44. Breiman L. Random Forests. Machine Learning. 2001;45: 5–32. doi:10.1023/A:1010933404324

45. Touw WG, Bayjanov JR, Overmars L, Backus L, Boekhorst J, Wels M, et al. Data mining in the Life Sciences with Random Forest: a walk in the park or lost in the jungle? Brief Bioinform. 2013;14: 315–326. doi:10.1093/bib/bbs034

46. Verikas A, Gelzinis A, Bacauskiene M. Mining data with random forests: A survey and results of new tests. Pattern Recognition. 2011;44: 330–349. doi:10.1016/j.patcog.2010.08.011

47. Wilcoxon F. Individual Comparisons by Ranking Methods. Biometrics Bulletin. 1945;1: 80–83. doi:10.2307/3001968

48. Benjamini Y, Hochberg Y. Controlling the False Discovery Rate: A Practical and Powerful Approach to Multiple Testing. Journal of the Royal Statistical Society Series B (Methodological). 1995;57: 289–300.

49. Rosenthal R. Parametric measures of effect size. The handbook of research synthesis. New York, NY, US: Russell Sage Foundation; 1994. pp. 231–244.

50. Appropriate statistics for ordinal level data: Should we really be using t-test and Cohen’sd for evaluating group differences on the NSSE and other surveys? | BibSonomy. [cited 12 Oct 2020]. Available: <<https://www.bibsonomy.org/bibtex/216a5c27e770147e5796719fc6b68547d/kweiand>>

