## Supplementary Information for "Metabolomic/lipidomic profiling of COVID-19 and individual response to tocilizumab"

**Supporting information**

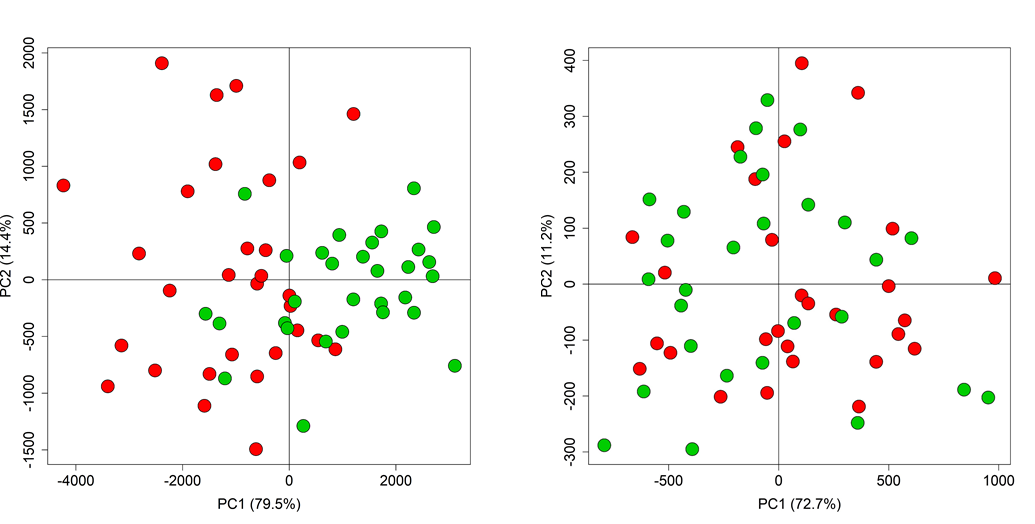

**S1 Fig.** **PCA analysis**. Score plots (PC1 vs. PC2) of the unsupervised PCA model of A) 21 quantified metabolites, B) lipoprotein-related parameters; COVID-19 patients (red dots); CTR subjects (green dots).

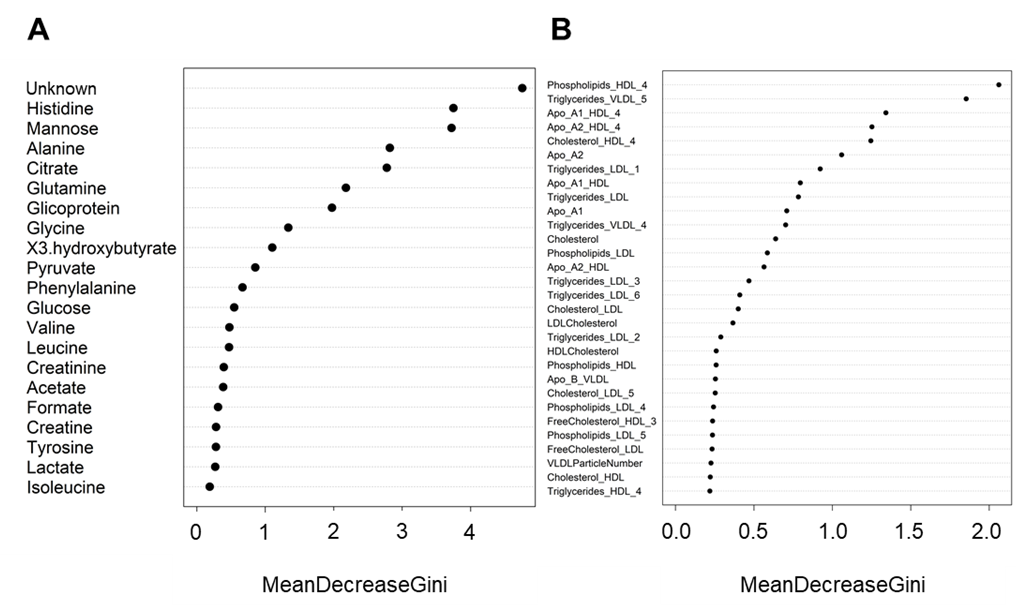

**S2 Fig.** **Metabolomic/lipidomic alterations in COVID-19 patients.** Variable importance plots of the Random Forest models discriminating COVID-19 patients and control subjects. A) 21 quantified metabolites, B) lipoprotein-related parameters.

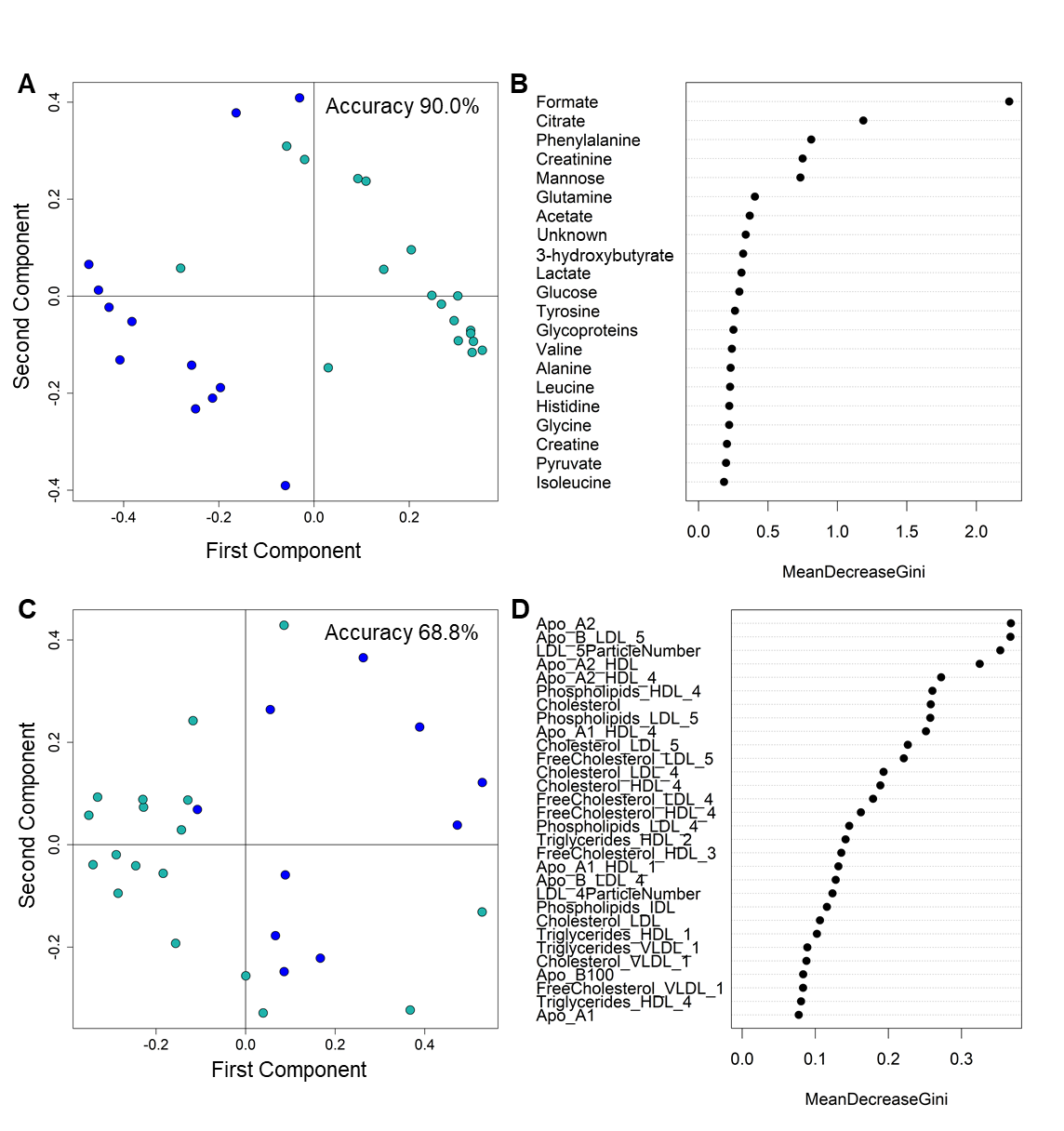
**S3** **Fig. Alterations induced by invasive ventilation in COVID-19 patients.** Proximity plot (of the first two principal components) and accuracy of the Random Forest model discriminating COVID-19 patients treated (blue dots) and non-treated (sea green dots) with invasive ventilation using metabolites (A) and lipoprotein-related parameters (C). Variable importance plots of the two Random Forest models: B) 21 quantified metabolites, D) lipoprotein-related parameters.

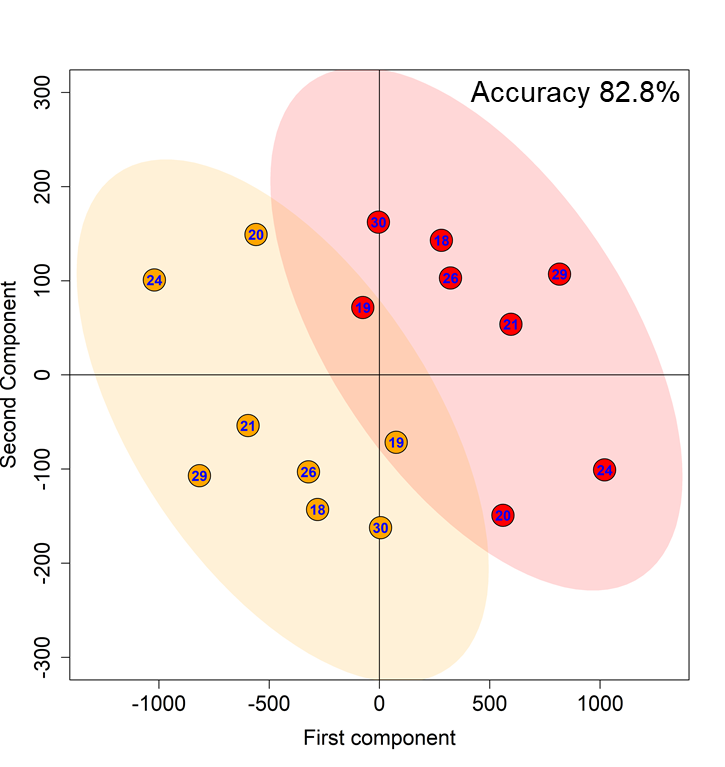

**S4 Fig.** **Alterations in lipoprotein profile induced by Tocilizumab treatment.** A) Score plot (of the first two principal components) and accuracy of the mPLS-DA model discriminating COVID-19 patients pre- (red dots) and post- (orange dots) tocilizumab treatment using the lipoprotein-related parameters.

**S1 Table**. **Demographic and clinical characteristics of COVID-19 patients.** ACF denotes acute cardiac failure, AI autoimmune disease, AKI acute kidney injury, ARDS acute respiratory distress syndrome, CKD chronic kidney disease, CVD cardiovascular disease, D deceased, DBT type 2 diabetes, DH discharged home, DYS dyslipidemia, H hypertension, K cancer, LTFU lost to follow-up. In the age column, the age class is indicated; I: 35-40 years old; II: 41-59 years old; III: 60-79 years old; IV: 80-85 years old.

|  | **Gender** | **Age**  **(35-85)** | **Comorbidities** | **Complications** | **Invasive ventilation** | **Outcome** |
| --- | --- | --- | --- | --- | --- | --- |
| **COVID-19-001** | F | III | H | ARDS | No | DH |
| **COVID-19-002** | M | III | H | None | No | DH |
| **COVID-19-003** | M | III | H, DBT | None | No | DH |
| **COVID-19-004** | F | III | H | None | No | DH |
| **COVID-19-005** | M | II | H, CVD | None | No | DH |
| **COVID-19-006** | F | III | H | None | No | DH |
| **COVID-19-007** | F | I | K | None | No | DH |
| **COVID-19-008** | M | II | None | None | No | DH |
| **COVID-19-009** | M | VI | H, CVD, DBT | ARDS | No | D |
| **COVID-19-010** | F | III | None | None | No | DH |
| **COVID-19-011** | M | III | None | None | No | DH |
| **COVID-19-012** | F | III | H | None | No | DH |
| **COVID-19-013** | F | III | K, DYS | ARDS | No | DH |
| **COVID-19-014** | M | III | H | ARDS | No | DH |
| **COVID-19-015** | F | II | None | None | No | DH |
| **COVID-19-016** | M | II | AI | ARDS | No | DH |
| **COVID-19-017** | F | III | None | None | No | DH |
| **COVID-19-018** | M | III | H | ARDS | Yes | LTFU |
| **COVID-19-019** | F | III | H | ARDS | Yes | DH |
| **COVID-19-020** | F | IV | CVD, CKD, DBT | ARDS, ACF | Yes | D |
| **COVID-19-021** | M | III | H, DYS | ARDS | Yes | D |
| **COVID-19-022** | M | III | CVD, DBT, K | ARDS | Yes | D |
| **COVID-19-023** | M | IV | H, CVD | ARDS, AKI | Yes | DH |
| **COVID-19-024** | M | III | None | ARDS | Yes | DH |
| **COVID-19-025** | M | III | CVD | ARDS | Yes | DH |
| **COVID-19-026** | M | III | CVD | ARDS | Yes | DH |
| **COVID-19-027** | M | II | None | ARDS | Yes | DH |
| **COVID-19-028** | F | I | None | ARDS | No | DH |
| **COVID-19-029** | M | III | H, DYS | ARDS | Yes | DH |
| **COVID-19-030** | M | III | CVD, DBT, K | ARDS | Yes | DH |

**S2 Table.** **Metabolomic/lipidomic alterations in COVID-19 patients: multivariate analysis.** Random Forest scores of the model discriminating COVID-19 patients and controls using the 21 quantified metabolites and lipoprotein-related parameters. P: predicted class; S. numeric score (controls: 0<S< 0.5; COVID-19 patients: 0.5 < S< 1).

|  | **Metabolites** |  | **Lipoproteins** |  |
| --- | --- | --- | --- | --- |
|  | **P** | **S** | **P** | **S** |
| **COVID-19-001** | COVID-19 | 0.951 | COVID-19 | 0.935 |
| **COVID-19-002** | COVID-19 | 0.832 | COVID-19 | 0.571 |
| **COVID-19-003** | COVID-19 | 0.995 | COVID-19 | 0.918 |
| **COVID-19-004** | COVID-19 | 0.817 | CTR | 0.296 |
| **COVID-19-005** | CTR | 0.268 | COVID-19 | 0.610 |
| **COVID-19-006** | COVID-19 | 0.779 | CTR | 0.474 |
| **COVID-19-007** | COVID-19 | 0.693 | CTR | 0.488 |
| **COVID-19-008** | COVID-19 | 0.789 | COVID-19 | 0.854 |
| **COVID-19-009** | COVID-19 | 0.646 | COVID-19 | 0.919 |
| **COVID-19-010** | COVID-19 | 0.789 | COVID-19 | 0.508 |
| **COVID-19-011** | COVID-19 | 0.620 | COVID-19 | 0.889 |
| **COVID-19-012** | COVID-19 | 0.579 | CTR | 0.347 |
| **COVID-19-013** | COVID-19 | 0.930 | COVID-19 | 0.516 |
| **COVID-19-014** | COVID-19 | 0.864 | COVID-19 | 0.906 |
| **COVID-19-015** | COVID-19 | 0.537 | COVID-19 | 0.739 |
| **COVID-19-016** | COVID-19 | 0.976 | COVID-19 | 0.780 |
| **COVID-19-017** | CTR | 0.469 | COVID-19 | 0.866 |
| **COVID-19-018** | COVID-19 | 0.914 | COVID-19 | 0.963 |
| **COVID-19-019** | COVID-19 | 0.824 | COVID-19 | 0.948 |
| **COVID-19-020** | COVID-19 | 0.871 | COVID-19 | 0.794 |
| **COVID-19-021** | COVID-19 | 0.899 | COVID-19 | 0.873 |
| **COVID-19-022** | COVID-19 | 0.961 | COVID-19 | 0.781 |
| **COVID-19-023** | COVID-19 | 0.895 | COVID-19 | 0.829 |
| **COVID-19-024** | COVID-19 | 0.911 | COVID-19 | 0.812 |
| **COVID-19-025** | COVID-19 | 0.977 | / | / |
| **COVID-19-026** | COVID-19 | 0.926 | COVID-19 | 0.777 |
| **COVID-19-027** | COVID-19 | 0.947 | / | / |
| **COVID-19-028** | COVID-19 | 0.848 | COVID-19 | 0.840 |
| **COVID-19-029** | COVID-19 | 0.835 | COVID-19 | 0.908 |
| **COVID-19-030** | COVID-19 | 0.696 | COVID-19 | 0.901 |
| **CTR-01** | CTR | 0.164 | CTR | 0.290 |
| **CTR-02** | COVID-19 | 0.752 | COVID-19 | 0.767 |
| **CTR-03** | CTR | 0.125 | CTR | 0.331 |
| **CTR-04** | CTR | 0.172 | / | / |
| **CTR-05** | CTR | 0.102 | CTR | 0.140 |
| **CTR-06** | CTR | 0.089 | CTR | 0.214 |
| **CTR-07** | CTR | 0.041 | / | / |
| **CTR-08** | CTR | 0.101 | CTR | 0.190 |
| **CTR-09** | CTR | 0.116 | CTR | 0.032 |
| **CTR-10** | CTR | 0.017 | CTR | 0.016 |
| **CTR-11** | CTR | 0.131 | CTR | 0.154 |
| **CTR-12** | CTR | 0.003 | CTR | 0.034 |
| **CTR-13** | CTR | 0.108 | CTR | 0.271 |
| **CTR-14** | CTR | 0.056 | CTR | 0.158 |
| **CTR-15** | CTR | 0.114 | CTR | 0.091 |
| **CTR-16** | CTR | 0.267 | CTR | 0.315 |
| **CTR-17** | COVID-19 | 0.641 | COVID-19 | 0.665 |
| **CTR-18** | CTR | 0.124 | CTR | 0.395 |
| **CTR-19** | CTR | 0.106 | CTR | 0.171 |
| **CTR-20** | COVID-19 | 0.521 | COVID-19 | 0.707 |
| **CTR-21** | CTR | 0.029 | CTR | 0.412 |
| **CTR-22** | CTR | 0.023 | CTR | 0.078 |
| **CTR-23** | CTR | 0.183 | CTR | 0.233 |
| **CTR-24** | CTR | 0.345 | CTR | 0.054 |
| **CTR-25** | CTR | 0.044 | CTR | 0.106 |
| **CTR-26** | CTR | 0.256 | CTR | 0.401 |
| **CTR-27** | CTR | 0.202 | CTR | 0.221 |
| **CTR-28** | CTR | 0.054 | CTR | 0.171 |
| **CTR-29** | CTR | 0.060 | CTR | 0.091 |
| **CTR-30** | CTR | 0.357 | CTR | 0.417 |

**S3 Table.** **Metabolomic alterations in COVID-19 patients: univariate analysis.** Univariate analysis of the 21 quantified metabolites for the comparison between COVID-19 patients and control subjects. The median and MAD of each metabolite in the two groups are reported. The p-value of the univariate Wilcoxon-Mann-Whitney test together with the p-value calculated after false discovery rate correction and the effect size, using the Cliff’s delta formulation, were also reported for each metabolite.

|  | **COVID-19** | | **CTR** | |  |  |  |
| --- | --- | --- | --- | --- | --- | --- | --- |
|  | **Median** | **MAD** | **Median** | **MAD** | **P-value** | **P-value FDR** | **Effect Size** |
| Acetate | 158.852 | 29.154 | 187.748 | 45.932 | 0.040 | 0.070 | small |
| Alanine | 574.375 | 101.069 | 801.354 | 133.587 | 0.000 | 0.000 | large |
| Unknown | 974.598 | 315.484 | 574.130 | 92.622 | 0.000 | 0.000 | large |
| Citrate | 91.353 | 28.299 | 169.619 | 39.353 | 0.000 | 0.000 | large |
| Creatine | 17.149 | 25.426 | 41.153 | 43.950 | 0.164 | 0.215 | small |
| Creatinine | 143.676 | 53.297 | 125.449 | 23.288 | 0.273 | 0.337 | small |
| Formate | 26.806 | 9.398 | 29.388 | 10.753 | 0.314 | 0.366 | small |
| Glycoproteins | 9968.183 | 1161.134 | 7955.922 | 1587.599 | 0.000 | 0.000 | large |
| Glucose | 3020.490 | 977.844 | 2757.041 | 389.345 | 0.138 | 0.193 | small |
| Glutamine | 358.677 | 87.285 | 455.643 | 52.495 | 0.000 | 0.000 | large |
| Glycine | 369.329 | 73.603 | 487.016 | 77.864 | 0.000 | 0.000 | large |
| Histidine | 108.327 | 14.399 | 134.834 | 23.553 | 0.000 | 0.000 | large |
| Isoleucine | 86.956 | 16.727 | 70.813 | 21.090 | 0.116 | 0.174 | small |
| Lactate | 751.451 | 201.780 | 736.530 | 287.680 | 0.971 | 0.971 | negligible |
| Leucine | 237.254 | 54.036 | 194.362 | 39.904 | 0.085 | 0.137 | small |
| Mannose | 47.346 | 15.782 | 20.935 | 8.821 | 0.000 | 0.000 | large |
| Phenylalanine | 166.993 | 48.926 | 139.680 | 22.319 | 0.007 | 0.013 | medium |
| Pyruvate | 469.671 | 80.759 | 365.668 | 81.428 | 0.002 | 0.004 | medium |
| Tyrosine | 173.344 | 35.193 | 183.844 | 25.633 | 0.730 | 0.767 | negligible |
| Valine | 932.619 | 104.354 | 888.724 | 105.917 | 0.572 | 0.632 | negligible |
| 3-hydroxybutyrate | 68.250 | 67.022 | 21.012 | 14.767 | 0.000 | 0.000 | large |

**S4 Table.** **Lipidomic alterations in COVID-19 patients: univariate analysis.** Univariate analysis of the lipoprotein-related parameters for the comparison between COVID-19 patients and control subjects. The median and MAD of each parameter in the two groups are reported. The p-value of the univariate Wilcoxon-Mann-Whitney test together with the p-value calculated after false discovery rate correction and the effect size, using the Cliff’s delta formulation, were also reported for each parameter.

|  | **COVID-19** | | **CTR** | |  |  |  |
| --- | --- | --- | --- | --- | --- | --- | --- |
|  | **Median** | **MAD** | **Median** | **MAD** | **P-value** | **P-value FDR** | **Effect Size** |
| Tg | 138.080 | 46.272 | 109.190 | 45.442 | 0.009 | 0.027 | medium |
| Chol | 157.845 | 37.072 | 214.110 | 25.849 | 0.000 | 0.000 | large |
| LDLChol | 77.690 | 23.106 | 119.670 | 21.053 | 0.000 | 0.000 | large |
| HDLChol | 42.540 | 9.140 | 61.710 | 17.917 | 0.001 | 0.003 | large |
| Apo A1 | 105.250 | 20.475 | 147.200 | 33.833 | 0.000 | 0.000 | large |
| Apo A2 | 24.415 | 6.116 | 32.335 | 5.656 | 0.000 | 0.000 | large |
| Apo B100 | 87.050 | 17.902 | 89.745 | 20.445 | 0.586 | 0.661 | negligible |
| LDLChol/HDLChol | 1.740 | 0.600 | 1.895 | 0.660 | 0.787 | 0.807 | negligible |
| Apo A1/Apo B100 | 0.755 | 0.170 | 0.600 | 0.215 | 0.003 | 0.011 | medium |
| Total(Particle number) | 1582.760 | 325.564 | 1631.800 | 371.621 | 0.586 | 0.661 | negligible |
| VLDL(Particle number) | 218.715 | 79.104 | 148.495 | 69.601 | 0.000 | 0.001 | large |
| IDL(Particle number) | 108.820 | 68.415 | 82.075 | 21.550 | 0.264 | 0.371 | small |
| LDL(Particle number) | 1189.120 | 256.156 | 1405.145 | 208.787 | 0.020 | 0.049 | medium |
| LDL1 (Particle number) | 222.025 | 58.377 | 213.080 | 80.980 | 0.714 | 0.761 | negligible |
| LDL2 (Particle number) | 161.745 | 47.866 | 200.120 | 101.002 | 0.230 | 0.332 | small |
| LDL3 (Particle number) | 128.545 | 53.514 | 169.780 | 66.569 | 0.499 | 0.584 | negligible |
| LDL4 (Particle number) | 98.925 | 97.436 | 158.890 | 67.088 | 0.043 | 0.089 | small |
| LDL5 (Particle number) | 152.915 | 93.152 | 197.290 | 127.704 | 0.011 | 0.029 | medium |
| LDL6 (Particle number) | 359.685 | 111.225 | 400.830 | 107.266 | 0.306 | 0.401 | small |
| Tg-VLDL | 86.365 | 39.430 | 65.075 | 39.793 | 0.020 | 0.049 | medium |
| Tg-IDL | 9.610 | 7.435 | 8.945 | 6.701 | 0.301 | 0.401 | small |
| Tg-LDL | 28.805 | 12.039 | 19.510 | 5.159 | 0.000 | 0.002 | large |
| Tg-HDL | 10.385 | 4.692 | 10.515 | 4.211 | 0.781 | 0.807 | negligible |
| Chol-VLDL | 21.075 | 7.317 | 19.925 | 8.169 | 0.469 | 0.557 | negligible |
| Chol-IDL | 12.235 | 8.844 | 11.135 | 2.787 | 0.743 | 0.784 | negligible |
| Chol-LDL | 77.690 | 23.106 | 119.670 | 21.053 | 0.000 | 0.000 | large |
| Chol-HDL | 42.540 | 9.140 | 61.710 | 17.917 | 0.001 | 0.003 | large |
| FreeChol-VLDL | 10.445 | 3.944 | 9.205 | 3.106 | 0.145 | 0.236 | small |
| FreeChol-IDL | 3.235 | 2.802 | 3.325 | 0.882 | 0.793 | 0.807 | negligible |
| FreeChol-LDL | 24.735 | 6.694 | 34.565 | 6.034 | 0.002 | 0.008 | medium |
| FreeChol-HDL | 10.340 | 2.802 | 13.115 | 5.411 | 0.106 | 0.176 | small |
| Ph-VLDL | 21.485 | 6.931 | 20.510 | 8.006 | 0.306 | 0.401 | small |
| Ph-IDL | 4.545 | 4.025 | 6.155 | 1.749 | 0.050 | 0.093 | small |
| Ph-LDL | 48.810 | 12.647 | 68.635 | 8.584 | 0.000 | 0.001 | large |
| Ph-HDL | 61.210 | 15.738 | 77.865 | 26.939 | 0.001 | 0.003 | large |
| Apo A1-HDL | 105.505 | 21.683 | 149.330 | 32.143 | 0.000 | 0.000 | large |
| Apo A2-HDL | 25.870 | 5.812 | 33.060 | 4.915 | 0.000 | 0.000 | large |
| Apo B-VLDL | 12.030 | 4.351 | 8.170 | 3.833 | 0.000 | 0.001 | large |
| Apo B-IDL | 5.985 | 3.766 | 4.515 | 1.186 | 0.258 | 0.368 | small |
| Apo B-LDL | 65.400 | 14.092 | 77.280 | 11.483 | 0.020 | 0.049 | medium |
| Tg-VLDL1 | 39.765 | 22.076 | 27.120 | 24.018 | 0.041 | 0.087 | small |
| Tg-VLDL2 | 15.295 | 5.545 | 9.320 | 4.618 | 0.009 | 0.026 | medium |
| Tg-VLDL3 | 15.015 | 3.543 | 9.990 | 3.966 | 0.008 | 0.025 | medium |
| Tg-VLDL4 | 13.595 | 6.123 | 8.855 | 2.513 | 0.000 | 0.000 | large |
| Tg-VLDL5 | 4.395 | 0.638 | 2.585 | 0.689 | 0.000 | 0.000 | large |
| Chol-VLDL1 | 5.770 | 2.973 | 5.205 | 2.758 | 1.000 | 1.000 | negligible |
| Chol-VLDL2 | 2.615 | 1.053 | 2.905 | 1.631 | 0.408 | 0.495 | negligible |
| Chol-VLDL3 | 3.495 | 1.423 | 3.675 | 1.824 | 0.712 | 0.761 | negligible |
| Chol-VLDL4 | 5.445 | 3.380 | 5.160 | 1.542 | 0.799 | 0.807 | negligible |
| Chol-VLDL5 | 1.650 | 0.786 | 1.600 | 0.808 | 0.204 | 0.306 | small |
| FreeChol-VLDL1 | 1.620 | 2.061 | 1.380 | 1.601 | 0.600 | 0.670 | negligible |
| FreeChol-VLDL2 | 1.705 | 0.697 | 1.055 | 0.504 | 0.014 | 0.037 | medium |
| FreeChol-VLDL3 | 1.995 | 0.697 | 1.385 | 0.667 | 0.071 | 0.127 | small |
| FreeChol-VLDL4 | 2.290 | 1.579 | 2.040 | 0.919 | 0.047 | 0.092 | small |
| FreeChol-VLDL5 | 0.825 | 0.408 | 0.910 | 0.408 | 0.781 | 0.807 | negligible |
| Ph-VLDL1 | 5.930 | 3.010 | 4.080 | 3.425 | 0.359 | 0.449 | negligible |
| Ph-VLDL2 | 3.380 | 1.475 | 2.615 | 1.001 | 0.213 | 0.315 | small |
| Ph-VLDL3 | 4.205 | 1.164 | 3.510 | 1.772 | 0.171 | 0.271 | small |
| Ph-VLDL4 | 5.880 | 2.802 | 4.550 | 1.379 | 0.013 | 0.036 | medium |
| Ph-VLDL5 | 2.000 | 0.719 | 1.730 | 0.815 | 0.062 | 0.112 | small |
| Tg-LDL1 | 9.725 | 5.130 | 6.015 | 1.164 | 0.000 | 0.000 | large |
| Tg-LDL2 | 3.425 | 1.646 | 2.235 | 0.875 | 0.001 | 0.003 | large |
| Tg-LDL3 | 3.615 | 1.090 | 2.770 | 0.675 | 0.000 | 0.002 | large |
| Tg-LDL4 | 2.780 | 1.727 | 2.030 | 1.001 | 0.021 | 0.050 | medium |
| Tg-LDL5 | 3.020 | 1.460 | 2.580 | 1.342 | 0.436 | 0.524 | negligible |
| Tg-LDL6 | 6.115 | 1.053 | 4.900 | 1.245 | 0.000 | 0.002 | large |
| Chol-LDL1 | 17.165 | 6.857 | 22.605 | 9.415 | 0.018 | 0.046 | medium |
| Chol-LDL2 | 12.525 | 5.352 | 19.570 | 12.105 | 0.049 | 0.093 | small |
| Chol-LDL3 | 8.420 | 4.959 | 15.825 | 6.397 | 0.040 | 0.087 | small |
| Chol-LDL4 | 6.015 | 8.918 | 13.480 | 5.130 | 0.002 | 0.008 | large |
| Chol-LDL5 | 9.255 | 6.724 | 15.405 | 10.423 | 0.001 | 0.005 | large |
| Chol-LDL6 | 23.280 | 8.288 | 25.605 | 9.622 | 0.276 | 0.383 | small |
| FreeChol-LDL1 | 5.955 | 1.749 | 6.690 | 3.025 | 0.106 | 0.176 | small |
| FreeChol-LDL2 | 4.805 | 1.475 | 6.345 | 3.677 | 0.346 | 0.443 | small |
| FreeChol-LDL3 | 3.880 | 1.601 | 5.630 | 2.068 | 0.390 | 0.478 | negligible |
| FreeChol-LDL4 | 2.870 | 1.809 | 4.215 | 1.327 | 0.044 | 0.089 | small |
| FreeChol-LDL5 | 3.405 | 1.623 | 4.605 | 2.254 | 0.005 | 0.017 | medium |
| FreeChol-LDL6 | 5.160 | 2.313 | 6.045 | 2.239 | 0.045 | 0.089 | small |
| Ph-LDL1 | 11.630 | 3.284 | 12.710 | 4.767 | 0.201 | 0.306 | small |
| Ph-LDL2 | 8.035 | 2.906 | 11.135 | 5.841 | 0.105 | 0.176 | small |
| Ph-LDL3 | 5.660 | 2.246 | 9.180 | 3.529 | 0.084 | 0.145 | small |
| Ph-LDL4 | 4.400 | 4.944 | 7.755 | 2.810 | 0.009 | 0.026 | medium |
| Ph-LDL5 | 5.770 | 3.677 | 8.515 | 4.944 | 0.002 | 0.008 | large |
| Ph-LDL6 | 13.020 | 3.684 | 14.000 | 4.715 | 0.193 | 0.301 | small |
| Apo B-LDL1 | 12.210 | 3.210 | 11.720 | 4.455 | 0.714 | 0.761 | negligible |
| Apo B-LDL2 | 8.895 | 2.639 | 11.005 | 5.552 | 0.230 | 0.332 | small |
| Apo B-LDL3 | 7.070 | 2.943 | 9.340 | 3.662 | 0.502 | 0.584 | negligible |
| Apo B-LDL4 | 5.440 | 5.360 | 8.740 | 3.684 | 0.043 | 0.089 | small |
| Apo B-LDL5 | 8.410 | 5.130 | 10.850 | 7.020 | 0.011 | 0.029 | medium |
| Apo B-LDL6 | 19.780 | 6.116 | 22.045 | 5.901 | 0.302 | 0.401 | small |
| Tg-HDL1 | 3.455 | 1.883 | 3.380 | 2.343 | 0.314 | 0.406 | small |
| Tg-HDL2 | 1.950 | 0.867 | 1.650 | 0.689 | 0.058 | 0.107 | small |
| Tg-HDL3 | 2.080 | 0.875 | 2.025 | 0.815 | 0.359 | 0.449 | negligible |
| Tg-HDL4 | 3.110 | 0.890 | 3.740 | 0.838 | 0.022 | 0.050 | medium |
| Chol-HDL1 | 12.625 | 4.796 | 17.365 | 11.549 | 0.523 | 0.602 | negligible |
| Chol-HDL2 | 6.625 | 2.810 | 7.790 | 3.647 | 0.198 | 0.306 | small |
| Chol-HDL3 | 7.670 | 2.276 | 9.450 | 3.017 | 0.005 | 0.017 | medium |
| Chol-HDL4 | 13.975 | 4.848 | 22.500 | 3.707 | 0.000 | 0.000 | large |
| FreeChol-HDL1 | 2.415 | 1.334 | 4.310 | 3.106 | 0.039 | 0.087 | small |
| FreeChol-HDL2 | 1.370 | 0.786 | 1.550 | 0.979 | 0.298 | 0.401 | small |
| FreeChol-HDL3 | 1.250 | 0.638 | 2.050 | 1.090 | 0.001 | 0.004 | large |
| FreeChol-HDL4 | 2.440 | 1.505 | 3.950 | 0.986 | 0.001 | 0.003 | large |
| Ph-HDL1 | 14.705 | 5.975 | 18.930 | 14.604 | 0.620 | 0.686 | negligible |
| Ph-HDL2 | 11.195 | 4.144 | 11.295 | 5.278 | 0.678 | 0.744 | negligible |
| Ph-HDL3 | 12.045 | 3.306 | 16.000 | 5.686 | 0.005 | 0.017 | medium |
| Ph-HDL4 | 19.235 | 4.789 | 29.520 | 4.522 | 0.000 | 0.000 | large |
| Apo A1-HDL1 | 13.440 | 7.183 | 23.065 | 18.088 | 0.075 | 0.132 | small |
| Apo A1-HDL2 | 14.460 | 4.270 | 17.800 | 6.968 | 0.008 | 0.025 | medium |
| Apo A1-HDL3 | 21.270 | 5.315 | 26.115 | 8.073 | 0.019 | 0.047 | medium |
| Apo A1-HDL4 | 54.920 | 15.627 | 82.065 | 8.733 | 0.000 | 0.000 | large |
| Apo A2-HDL1 | 1.680 | 1.112 | 2.285 | 1.757 | 0.372 | 0.461 | negligible |
| Apo A2-HDL2 | 2.370 | 0.956 | 2.970 | 1.379 | 0.147 | 0.236 | small |
| Apo A2-HDL3 | 4.625 | 1.638 | 6.015 | 1.564 | 0.046 | 0.090 | small |
| Apo A2-HDL4 | 13.685 | 5.552 | 20.815 | 3.417 | 0.000 | 0.000 | large |

**S5 Table. Metabolomic alterations induced by Tocilizumab treatment: univariate analysis.** Univariate analysis of the 21 quantified metabolites for the comparison between COVID-19 patients before and after tocilizumab treatment. The p-value of the univariate Wilcoxon-Signed-Rack test together with the p-value calculated after false discovery rate correction and the effect size were reported for each metabolite.

|  | **P-value** | **P-value FDR** | **Effect Size** | **Post-treatment trend** |
| --- | --- | --- | --- | --- |
| Leucine | 1.48E-01 | 2.60E-01 | small | ↓ |
| Isoleucine | 3.13E-01 | 5.05E-01 | small | ↑ |
| Valine | 9.45E-01 | 9.45E-01 | negligible | ↑ |
| Alanine | 7.81E-02 | 1.82E-01 | medium | ↑ |
| Acetate | 3.91E-02 | 1.03E-01 | medium | ↑ |
| Glycoproteins | 6.41E-01 | 7.47E-01 | negligible | ↑ |
| 3-hydroxybutyrate | 9.45E-01 | 9.45E-01 | negligible | ↓ |
| Pyruvate | 1.56E-02 | 8.20E-02 | medium | ↓ |
| Glutamine | 1.56E-02 | 8.20E-02 | medium | ↑ |
| Citrate | 7.81E-03 | 8.20E-02 | medium | ↑ |
| Glycine | 7.81E-03 | 8.20E-02 | medium | ↑ |
| Creatine | 1.48E-01 | 2.60E-01 | small | ↑ |
| Creatinine | 1.09E-01 | 2.30E-01 | small | ↑ |
| Lactate | 2.34E-02 | 8.20E-02 | medium | ↓ |
| Mannose | 2.34E-02 | 8.20E-02 | medium | ↓ |
| Glucose | 7.42E-01 | 8.20E-01 | negligible | ↑ |
| Tyrosine | 6.41E-01 | 7.47E-01 | negligible | ↑ |
| Histidine | 5.47E-01 | 7.47E-01 | negligible | ↑ |
| Unknown | 6.41E-01 | 7.47E-01 | negligible | ↓ |
| Phenylalanine | 3.91E-02 | 1.03E-01 | medium | ↓ |
| Formate | 3.83E-01 | 5.74E-01 | negligible | ↓ |

**S6 Table.** **Lipidomic alterations induced by Tocilizumab treatment: univariate analysis.** Univariate analysis of the lipoprotein-related parameters for the comparison between COVID-19 patients before and after tocilizumab treatment. The p-value of the univariate Wilcoxon-Signed-Rack test together with the p-value calculated after false discovery rate correction and the effect size were reported for each parameter.

|  | **P-value** | **P-value FDR** | **Effect Size** | **Post-treatment trend** |
| --- | --- | --- | --- | --- |
| Tg | 0.008 | 0.047 | large | ↑ |
| Chol | 0.055 | 0.107 | medium | ↑ |
| LDLChol | 0.250 | 0.320 | small | ↑ |
| HDLChol | 0.383 | 0.441 | small | ↓ |
| Apo A1 | 0.945 | 0.954 | negligible | ↓ |
| Apo A2 | 0.250 | 0.320 | small | ↑ |
| Apo B100 | 0.039 | 0.084 | medium | ↑ |
| LDLChol/HDLChol | 0.109 | 0.178 | medium | ↑ |
| Apo A1/Apo B100 | 0.078 | 0.141 | medium | ↑ |
| Total(Particle number) | 0.039 | 0.084 | medium | ↑ |
| VLDL(Particle number) | 0.016 | 0.056 | large | ↑ |
| IDL(Particle number) | 0.039 | 0.084 | medium | ↑ |
| LDL(Particle number) | 0.078 | 0.141 | medium | ↑ |
| LDL1 (Particle number) | 0.195 | 0.278 | small | ↑ |
| LDL2 (Particle number) | 0.742 | 0.783 | negligible | ↓ |
| LDL3 (Particle number) | 0.945 | 0.954 | negligible | ↑ |
| LDL4 (Particle number) | 0.108 | 0.178 | medium | ↑ |
| LDL5 (Particle number) | 0.008 | 0.047 | large | ↑ |
| LDL6 (Particle number) | 0.205 | 0.285 | small | ↑ |
| Tg-VLDL | 0.008 | 0.047 | large | ↑ |
| Tg-IDL | 0.008 | 0.047 | large | ↑ |
| Tg-LDL | 0.078 | 0.141 | medium | ↑ |
| Tg-HDL | 0.008 | 0.047 | large | ↑ |
| Chol-VLDL | 0.016 | 0.056 | large | ↑ |
| Chol-IDL | 0.016 | 0.056 | large | ↑ |
| Chol-LDL | 0.250 | 0.320 | small | ↑ |
| Chol-HDL | 0.383 | 0.441 | small | ↓ |
| FreeChol-VLDL | 0.016 | 0.056 | large | ↑ |
| FreeChol-IDL | 0.008 | 0.047 | large | ↑ |
| FreeChol-LDL | 0.148 | 0.238 | small | ↑ |
| FreeChol-HDL | 0.461 | 0.515 | small | ↓ |
| Ph-VLDL | 0.016 | 0.056 | large | ↑ |
| Ph-IDL | 0.008 | 0.047 | large | ↑ |
| Ph-LDL | 0.109 | 0.178 | medium | ↑ |
| Ph-HDL | 0.945 | 0.954 | negligible | ↓ |
| Apo A1-HDL | 0.945 | 0.954 | negligible | ↑ |
| Apo A2-HDL | 0.078 | 0.141 | medium | ↑ |
| Apo B-VLDL | 0.016 | 0.056 | large | ↑ |
| Apo B-IDL | 0.039 | 0.084 | medium | ↑ |
| Apo B-LDL | 0.078 | 0.141 | medium | ↑ |
| Tg-VLDL1 | 0.008 | 0.047 | large | ↑ |
| Tg-VLDL2 | 0.008 | 0.047 | large | ↑ |
| Tg-VLDL3 | 0.039 | 0.084 | medium | ↑ |
| Tg-VLDL4 | 0.023 | 0.069 | medium | ↑ |
| Tg-VLDL5 | 0.039 | 0.084 | medium | ↑ |
| Chol-VLDL1 | 0.008 | 0.047 | large | ↑ |
| Chol-VLDL2 | 0.016 | 0.056 | large | ↑ |
| Chol-VLDL3 | 0.039 | 0.084 | medium | ↑ |
| Chol-VLDL4 | 0.039 | 0.084 | medium | ↑ |
| Chol-VLDL5 | 0.055 | 0.107 | medium | ↑ |
| FreeChol-VLDL1 | 0.008 | 0.047 | large | ↑ |
| FreeChol-VLDL2 | 0.016 | 0.056 | large | ↑ |
| FreeChol-VLDL3 | 0.023 | 0.069 | medium | ↑ |
| FreeChol-VLDL4 | 0.023 | 0.069 | medium | ↑ |
| FreeChol-VLDL5 | 0.016 | 0.056 | large | ↑ |
| Ph-VLDL1 | 0.008 | 0.047 | large | ↑ |
| Ph-VLDL2 | 0.008 | 0.047 | large | ↑ |
| Ph-VLDL3 | 0.039 | 0.084 | medium | ↑ |
| Ph-VLDL4 | 0.023 | 0.069 | medium | ↑ |
| Ph-VLDL5 | 0.055 | 0.107 | medium | ↑ |
| Tg-LDL1 | 0.039 | 0.084 | medium | ↑ |
| Tg-LDL2 | 0.250 | 0.320 | small | ↑ |
| Tg-LDL3 | 0.250 | 0.320 | small | ↑ |
| Tg-LDL4 | 0.055 | 0.107 | medium | ↑ |
| Tg-LDL5 | 0.039 | 0.084 | medium | ↑ |
| Tg-LDL6 | 0.313 | 0.387 | small | ↑ |
| Chol-LDL1 | 0.195 | 0.278 | small | ↑ |
| Chol-LDL2 | 0.195 | 0.278 | small | ↓ |
| Chol-LDL3 | 0.461 | 0.515 | small | ↓ |
| Chol-LDL4 | 0.178 | 0.277 | small | ↑ |
| Chol-LDL5 | 0.022 | 0.069 | medium | ↑ |
| Chol-LDL6 | 0.353 | 0.428 | small | ↑ |
| FreeChol-LDL1 | 0.313 | 0.387 | small | ↑ |
| FreeChol-LDL2 | 0.195 | 0.278 | small | ↓ |
| FreeChol-LDL3 | 0.641 | 0.696 | negligible | ↓ |
| FreeChol-LDL4 | 0.383 | 0.441 | small | ↑ |
| FreeChol-LDL5 | 0.039 | 0.084 | medium | ↑ |
| FreeChol-LDL6 | 0.353 | 0.428 | small | ↑ |
| Ph-LDL1 | 0.195 | 0.278 | small | ↑ |
| Ph-LDL2 | 0.250 | 0.320 | small | ↓ |
| Ph-LDL3 | 0.742 | 0.783 | negligible | ↓ |
| Ph-LDL4 | 0.151 | 0.239 | small | ↑ |
| Ph-LDL5 | 0.022 | 0.069 | medium | ↑ |
| Ph-LDL6 | 0.383 | 0.441 | small | ↑ |
| Apo B-LDL1 | 0.195 | 0.278 | small | ↑ |
| Apo B-LDL2 | 0.742 | 0.783 | negligible | ↓ |
| Apo B-LDL3 | 0.945 | 0.954 | negligible | ↑ |
| Apo B-LDL4 | 0.108 | 0.178 | medium | ↑ |
| Apo B-LDL5 | 0.008 | 0.047 | large | ↑ |
| Apo B-LDL6 | 0.205 | 0.285 | small | ↑ |
| Tg-HDL1 | 0.547 | 0.605 | negligible | ↑ |
| Tg-HDL2 | 0.092 | 0.165 | medium | ↑ |
| Tg-HDL3 | 0.016 | 0.056 | large | ↑ |
| Tg-HDL4 | 0.016 | 0.056 | large | ↑ |
| Chol-HDL1 | 0.008 | 0.047 | large | ↓ |
| Chol-HDL2 | 0.109 | 0.178 | medium | ↓ |
| Chol-HDL3 | 1.000 | 1.000 | negligible | ↓ |
| Chol-HDL4 | 0.016 | 0.056 | large | ↑ |
| FreeChol-HDL1 | 0.039 | 0.084 | medium | ↓ |
| FreeChol-HDL2 | 0.383 | 0.441 | small | ↓ |
| FreeChol-HDL3 | 0.641 | 0.696 | negligible | ↑ |
| FreeChol-HDL4 | 0.052 | 0.107 | medium | ↑ |
| Ph-HDL1 | 0.008 | 0.047 | large | ↓ |
| Ph-HDL2 | 0.250 | 0.320 | small | ↓ |
| Ph-HDL3 | 0.109 | 0.178 | medium | ↑ |
| Ph-HDL4 | 0.008 | 0.047 | large | ↑ |
| Apo A1-HDL1 | 0.016 | 0.056 | large | ↓ |
| Apo A1-HDL2 | 0.195 | 0.278 | small | ↓ |
| Apo A1-HDL3 | 0.461 | 0.515 | small | ↓ |
| Apo A1-HDL4 | 0.023 | 0.069 | medium | ↑ |
| Apo A2-HDL1 | 0.008 | 0.047 | large | ↓ |
| Apo A2-HDL2 | 0.313 | 0.387 | small | ↓ |
| Apo A2-HDL3 | 0.039 | 0.084 | medium | ↑ |
| Apo A2-HDL4 | 0.008 | 0.047 | large | ↑ |
